# A Large Scale Multi-dataset Investigation of Brain Metastases Distribution Based on Primary Cancer Type

**DOI:** 10.1101/2024.09.30.24314304

**Authors:** Qing Lyu, Hongyu Yuan, Zhen Lin, Richard Barcus, Jeremy Hudson, Yuming Jiang, Jeongchul Kim, Christopher T Whitlow

## Abstract

Brain metastases (BM) present a significant clinical challenge, characterized by poor prognosis and high morbidity. Previous research has demonstrated heterogeneous distribution patterns of BMs depending on the primary tumor type. This study aims to further investigate the distribution patterns of BMs. Utilizing three public datasets and one private dataset, we analyzed 1,344 patients with a total of 9,431 identified BMs, categorizing them into lung, breast, melanoma, kidney cancer, and other groups. Our findings indicate that lung cancer BMs predominantly localize in the cerebellum, as well as the frontal, temporal, occipital, and parietal lobes. Breast cancer BMs are commonly found in the cerebellum, frontal, occipital, and temporal lobes, while melanoma BMs are most frequently located in the occipital, temporal, and frontal lobes. Kidney cancer BMs, less studied in existing literature, primarily affect the frontal and temporal lobes. Additionally, we explored the relationship between BM volume and tumor type, revealing that lung and breast cancer BMs are more likely to have smaller tumor volumes compared to melanoma and kidney cancer BMs. These findings confirm and expand upon previous research, offering new insights into the distribution and characteristics of kidney cancer BMs.

## Introduction

Brain metastases (BM) are the most common malignant tumors affecting the central nervous system, occurring in 30-40% of cancer patients at some stage of their disease^1^. BMs arise when tumor cells from primary cancers elsewhere in the body metastasize to the brain. Depending on the origin of these tumor cells, BMs are categorized into several types, with lung cancer being the most frequent (39% - 56%), followed by breast cancer (13% - 30%) and melanoma (8% - 11%). Less common sources include gastrointestinal (6% - 9%) and kidney cancers (2% - 6%)^2^. The prognosis of patients with BM remains poor, with a median survival time of less than 12 months^3^. The National Comprehensive Cancer Network (NCCN) recommends a multimodal treatment approach, which may include whole brain radiotherapy, stereotactic radiosurgery, surgical resection, chemotherapy, immunotherapy, targeted therapy, and systemic therapy^1,4,5^. The choice of treatment is based on several factors, including the type of primary tumor, the number and size of BM, and the location of the brain lesions^6^.

Studies have shown that the intracranial distribution of BMs is heterogeneous and has an implicit relationship with the primary cancer type^6^. The “Seed and Soil” theory was developed to explain this phenomenon^7^. According to this theory, tumor cells, or “seeds”, metastasize to specific brain regions, or “soil”, because these regions provide a unique microenvironment conductive to the growth of tumor cells. Recent extensions of the “Seed and Soil” theory suggest that the primary cancer origin influences both the “seed” and the “soil” by affecting cellular receptor and protein expression^8-10^. Research indicates that the occipital lobe and cerebellum are common metastasis sites for breast and lung cancer BMs, while frontal lobes are more commonly affected in melanoma BMs. However, there is a lack of studies exploring the distribution patterns of less common BM types.

In this study, we aim to investigate the intracranial distribution of BMs based on their primary cancer origins. Three publicly available dataset and a private dataset will be used, encompassing a total of 1,420 patients with 9,266 identified brain metastases. Our findings on common BM types, such as those originating from lung, breast, and melanoma cancers, will be compared with existing studies to enhance the current understanding of their distribution patterns. Additionally, our research into the intracranial distribution of less common BM types, such as those from kidney cancer, will help address existing gaps in knowledge on this topic.

### Related studies

In recent years, several studies have focused on investigating the location of BMs based on their primary cancer origins and analyzing factors that may influence their anatomical distribution. However, most of these studies have concentrated on common types, such as lung cancer, breast, and melanoma, with only a few providing insights into the distribution of less common BM types^6^.

#### Lung cancer BM distribution

As the most common type of BM, the distribution of lung cancer BM has been investigated by several research groups. Bender *et al*. studied 85 lung cancer patients and found that lung cancer metastasized to the cerebellum more frequently than expected, based on the relative brain volume^11^. Similarly, Quattrocchi *et al*. analyzed 57 patients and observed a higher likelihood of metastasis in the occipital lobes and cerebellum^12^. Takano *et al*., in a study involving 200 patients, revealed that lung cancer BMs commonly occurred in the cerebellum, occipital, and temporal lobes^13^. Wang *et al*. examined 335 lung cancer patients and found that the cerebellum, right parietal lobe, and frontal lobes had the highest rates of metastases^14^. Schroeder *et al*., after studying 167 patients, concluded that lung cancer BMs were more likely to occur in infratentorial areas^2^. Neman *et al*., in an analysis of data from 226 patients, revealed that lung cancer BMs were more likely to be located in the temporal lobes^15^. In contrast, Mampre *et al*. after investigating 195 patients, found no predilection for lung cancer BMs to metastasize to any particular vascular region^16^.

#### Breast cancer BM distribution

Breast cancer BMs have been investigated by multiple research groups, with several findings highlighting the cerebellum as a frequent metastasis site. Bender *et al*., after studying 30 patients, found that breast cancer was more like to metastasize to the cerebellum^11^. Quattrocchi *et al*. reported similar results in their study of 26 patients^*12*^. Kyeong *et al*. investigated breast cancer subtypes and found no significant differences in the number of BMs across subtypes, although some showed a predilection for the occipital, temporal, frontal lobes, and cerebellum^17^. Laakmann *et al*. also studied breast cancer subtypes and concluded that HER2+ patients had a higher incidence of cerebellar metastases compared to HER2-patients, while ER+ and PR+ patients had a lower incidence of hippocampal metastases^*18*^. Mampre *et al*., in a study of 75 patients, found that breast cancer BMs were less likely to occur in posterior cerebral artery territories and more likely to appear in interior-posterior watershed regions^16^. Schroeder *et al* analyzed data from 47 patients and concluded that breast cancer metastases favored areas of posterior circulation, being more likely located in the cerebellum and less likely in the frontal lobes^2^. Neman *et al*. found that breast cancer was more likely to metastasize to the right cerebellar hemisphere^*15*^. Young et al. investigated 34 patients and found breast cancer BMs emerged in the cerebellum, frontal, occipital, temporal, and parietal lobes^19^.

#### Melanoma BM distribution

Melanoma BMs have been studied with varying results regarding their preferred locations in the brain. One study involving 29 patients found that melanoma was no more likely to metastasize to the cerebellum than other brain regions^11^. Rogne *et al*. reported similar findings^20^. Another study, conducted on 56 patients, concluded that melanoma BMs were more likely to be located in the lateral lenticulostriate and medial lenticulostriate and less likely in the cerebellar vascular territory^16^. Neman *et al*., after investigating 483 patients, found that melanoma metastases were most commonly located in the left temporal lobe^15^. Schroeder *et al*. suggested that the frontal lobes were more likely to host melanoma metastases, while the cerebellum was not a frequent site^2^.

#### Kidney cancer BM distribution

There are limited studies on the distribution of BMs from kidney cancer. Seidel *et al*., in a study of 17 patients, found that renal cell carcinoma metastases were more likely to be in deep white matter^9^. Another study involving 89 patients suggested that the brainstem was a common site for kidney cancer BMs^15^. However, a separate study analyzing 44 metastases concluded that there was no specific predilection for any particular cerebral vascular territory^16^.

### Methodology

#### Dataset

This study investigated four datasets: one from Wake Forest Baptist Medical Center (WFBMC) and three publicly available datasets from the University of California - San Francisco, Universidad de Castilla - La Mancha, and Stanford University. Table I provides detailed information on the data used. There are 1,344 patients involved into this study, with 9,431 identified brain metastases. Among these patients, 748 (56%) of them belong to the lung cancer type, 212 (16%) belong to the breast cancer type, 193 (14%) belong to the melanoma type, 77 (6%) belong to the kidney cancer type, and 114 (8%) belong to other types.

##### WFBMC Dataset

In this study, we collected 812 patients who underwent gamma knife treatment in WFBMC during 2005 and 2021. Among these patients, 148 of them are with BM masks labeled by radiologists and all the other patients are without BM masks. Images were collected using GE 1.5T Signa HDxt, GE 3.0T Discovery MR750 scanners, GE 3T Signa Areo, and Siemens 3T Skyra scanners. T1 contrast-enhanced images were collected. The dataset used in this study was collected under Wake Forest University School of Medicine Institutional Review Board approval (IRB00002960) and in accordance with ethical guidelines ensuring patient privacy and confidentiality.

##### UCSF dataset

The University of California, San Francisco Brain Metastases Stereotactic Radiosurgery (UCSF-BMSR) MRI Dataset includes data from 412 patients who underwent gamma knife radiosurgery between 2017 and 2020^21^. The dataset contains registered and skull-stripped multi-modality MRI images with segmented BM annotations, featuring T1 post-contrast, T1 pre-contrast, Fluid attenuated inversion recovery (FLAIR), and subtracted images of T1 pre- and post-contrast images. Data was acquired using GE 1.5T Signa HDxt, Philips 1.5T Achieva, and GE 3.0T Discovery MR750 scanners. For this study, the training subset was utilized, consisting of 314 patients and a total of 1,926 BMs. Patients were grouped into 22 categories based on the primary cancer type. To ensure consistency with other datasets, we retained the lung, breast, melanoma, and kidney categories, while merging the remaining groups under the “other” category.

##### Molab dataset

The Universidad de Castilla - La Mancha, Mathematical Oncology Laboratory (Molab) BM dataset consists of 75 patients with 260 BM lesions^22^. A total of 593 imaging sequences were collected from 5 different medical institutions between 2005 and 2021. The dataset features multimodal MRI scans, including T1 pre-contrast, T1 post-contrast, T2, and FLAIR. BMs were segmented using an in-house semi-automatic segmentation procedure^23,24^. Patients were classified by cancer type, with categories other than lung, breast, kidney, and melanoma grouped under “other” to align with other datasets. Since multiple imaging sequences were available for each patient, this study utilized only the first MRI scan for each patient with corresponding segmented BM annotations. The dataset provides two segmentation masks for each BM: one for contrast-enhancing regions and another for the non-enhancing or necrotic areas. The union of these two masks was selected as the tumor ground truth.

##### Standford dataset

The Standford BrainMetShare dataset consists of 156 patients, collected between 2016 and 2018^25^. Multi-modality MRI images, including T1 pre-contrast, T1 post-contrast, and FLAIR, were collected using GE 1.5T Signa Explorer, GE 3T Discovery MR750, GE 3T Discovery MR750w, GE 3T Signa Architect, and Siemens 3T Skyra scanners. BM segmentation was performed by two neuroradiologists. For this study, after excluding patients without a confirmed cancer type, 144 patients were included. Patients were categorized by cancer type, with categories other than lung, breast, kidney, and melanoma grouped under “other” to maintain consistency with other datasets.

#### Data preprocessing

We employed the ANTs nonlinear registration tool (https://github.com/ANTsX/ANTs) to align all datasets, including both brain images and metastases segmentation masks, to the MNI152 1mm template^26^. All transformed segmentation masks were preserved, despite any intensity variations caused by interpolation during the registration process. We explored several existing brain extraction algorithms, including the FreeSurfer^27^, FMRIB Software Library (FSL)^28^, HD-BET (https://github.com/MIC-DKFZ/HD-BET)^29^, and nnUNet^30^. After evaluation, we selected nnUNet for its effectiveness in isolating the brain region.

#### Brain metastases segmentation

We used nnUNet to segment BM for the scans without human labeled BM contours within the WFBMC dataset^30^. For the 148 patients with BM masks in the WFBMC dataset, we randomly selected 130 patients and combined them with all patients in Molab, UCSF, and Standford datasets to create a training dataset. The remaining 18 patients with BM masks within WFBMC dataset were utilized for validation. According to our results on the validation dataset, nnUNet achieved an overall Dice score of 0.92 with a sensitivity of 0.89 and specificity of 0.93. Figure 1 shows examples of our segmentation results on unsegmented WFBMC dataset. After training, our utilized the trained nnUNet to segment all unsegmented patients in the WFBMC dataset. All the training, validation, and testing experiments were conducted using Pytorch on an Nvidia A6000 GPU.

**Figure 1.**
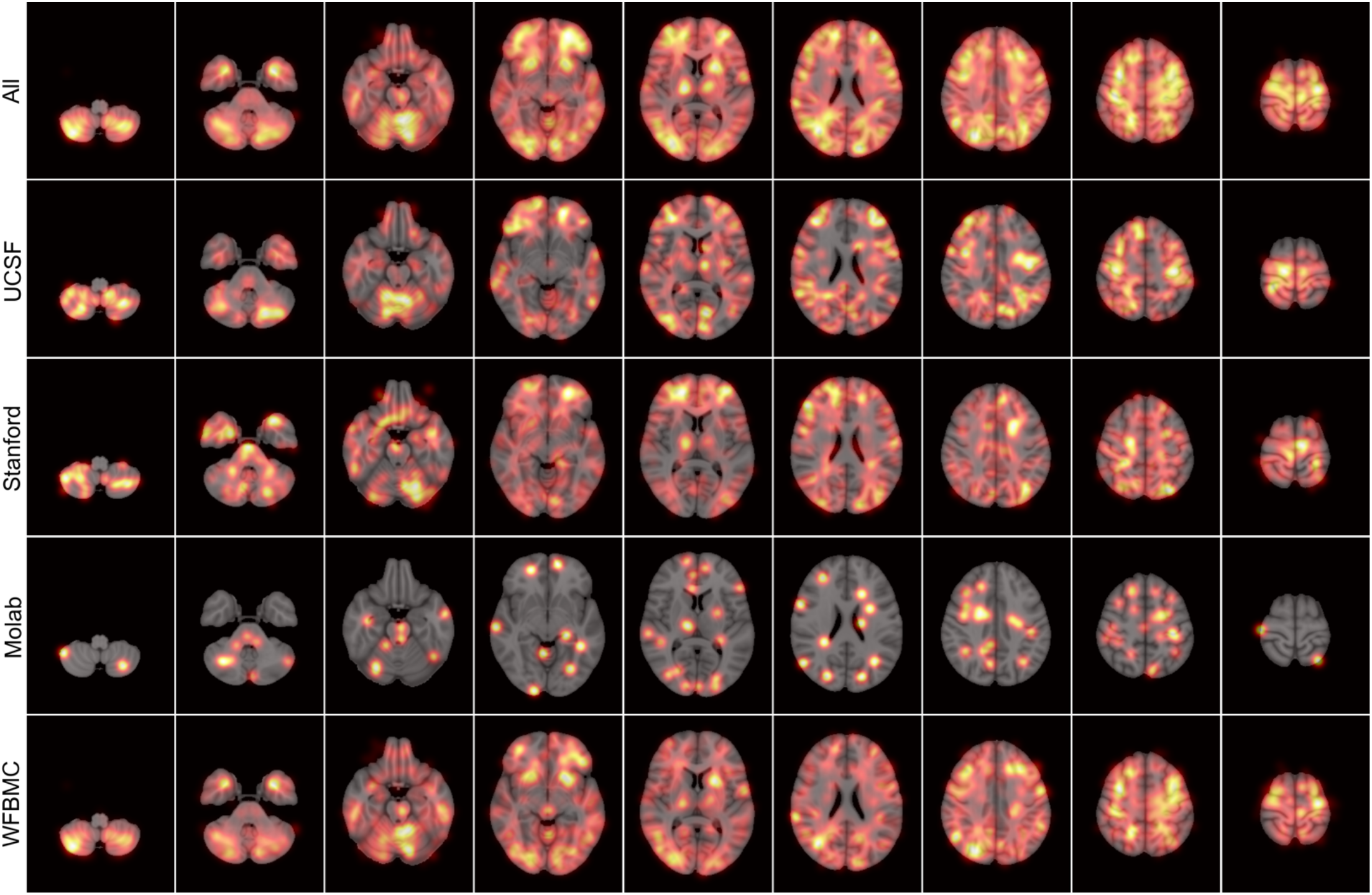
Lung cancer BM frequency distribution maps.

#### BM centroid computation

We utilized the Python Scipy library to calculate the centroids of BMs^31^. The coordinates of these BM centroids were recorded and used to identify their corresponding regions on two brain atlas templates: the Automated Anatomical Labeling (AAL)^32^ template and the Artery atlas^33^ template. Additionally, we developed a custom atlas, termed the AAL Group atlas, based on the AAL template. This new atlas categorizes each AAL region into one of nine groups: Frontal, Occipital, Parietal, Cerebellum, Temporal, Caudate, Putamen, Pallidum, and Thalamus.

## Result

To study the distribution of BM, we created frequency distribution maps for each BM type and performed statistical analysis of BM centroid locations using the AAL, AAL Group, and Artery Atlas.

### Lung cancer BM distribution

Lung cancer BMs are the most common type observed, with a total of 4,878 BMs studied, representing 52% of all BMs analyzed in this study. Figure 1 shows frequency distribution maps based on lung cancer BMs across the UCSF, Stanford, Molab, and WFBMC datasets. The frequency distribution maps are drawn based on statistics of BM centroid locations in the adjacent slides of the demonstration axial slice. It can be found results from different datasets demonstrate consistent results that regions such as cerebellum, frontal, occipital, temporal, and parietal lobes are the most likely sites for lung cancer BMs.

We analyzed the distribution of BM centroids using the AAL and AAL Group atlases. Figure 2(a) illustrates the frequency of lung cancer BM centroids in specific regions of the AAL Group atlas, while Table 2 identifies the ten most common AAL regions where lung cancer BMs were observed. The frontal and temporal regions emerge as the most prevalent across all datasets, with the Stanford dataset showing a notable occurrence of BMs in the occipital region. According to Table 2, some frontal areas such as the Right Superior Frontal Gyrus, Left Middle Frontal Gyrus, and right Middle Frontal Gyrus are consistently identified in all four dataset results. Additionally, the Right Precentral Gyrus and Right Cerebellum (Crus 1) appear in three out of four dataset results. Other regions, including the Left Precentral Gyrus, Left Middle Occipital Gyrus, Left Postcentral Gyrus, Left Precuneus, and Right Middle Temporal Gyrus, are found in two of the four dataset results.

**Table 1.**
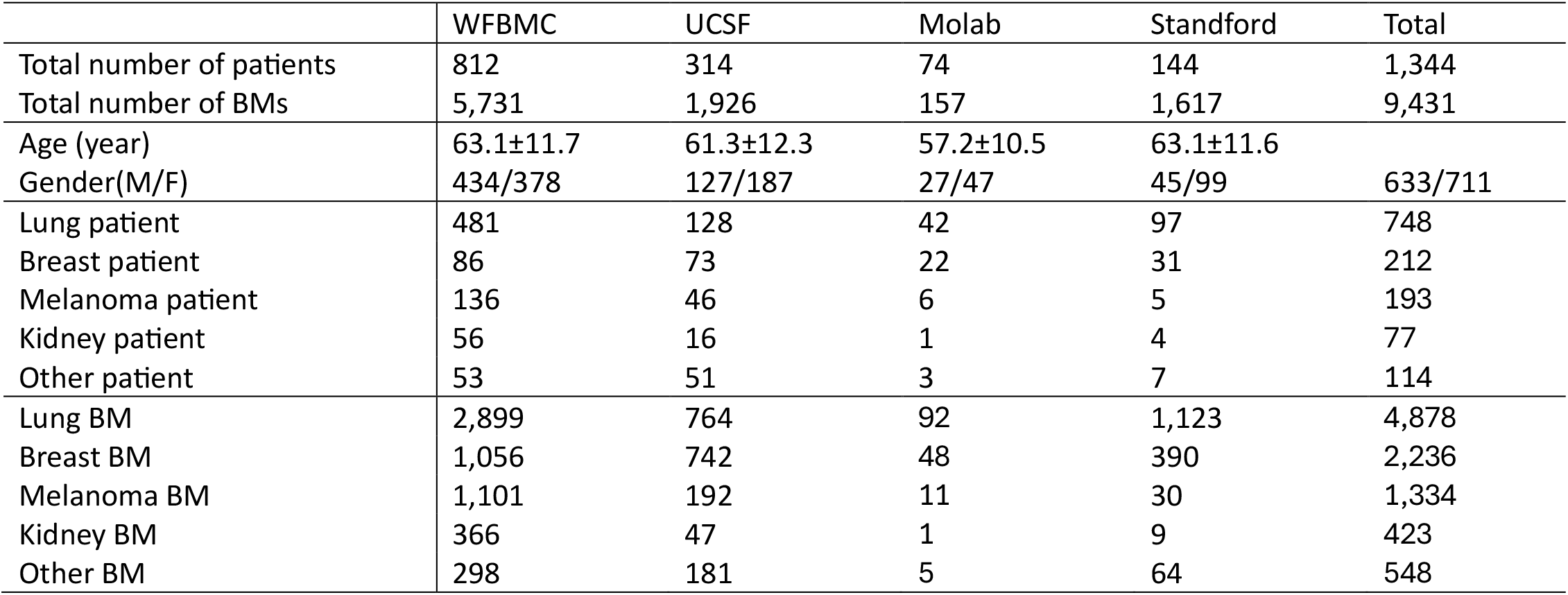
Summary of the four datasets used in this study.

**Table 2.** The ten most common AAL regions for lung cancer BMs.

|  | UCSF | Stanford | Molab | WFBMC |
| --- | --- | --- | --- | --- |
| 1<sup>st</sup> | Precentral_R | Occipital_Sup_R | Frontal_Sup_R | Precentral_L |
| 2<sup>nd</sup> | Frontal_Mid_R | Frontal_Mid_R | Precentral_R | Frontal_Mid_R |
| 3<sup>rd</sup> | Frontal_Mid_L | Frontal_Mid_L | Frontal_Mid_R | Cerebelm_Crus2_R |
| 4<sup>th</sup> | Temporal_Inf_L | Frontal_Sup_R | Cerebelm_Crus1_R | Frontal_Sup_R |
| 5<sup>th</sup> | Temporal_Mid_R | Frontal_Sup_L | Cerebelum_4_5_R | Frontal_Mid_L |
| 6<sup>th</sup> | Precentral_L | Front_Sup_Med_L | Frontal_Mid_L | Cerebelum_8_L |
| 7<sup>th</sup> | Parietal_Inf_L | Postcentral_L | Supp_Motor_L | Cerebelum_8_R |
| 8<sup>th</sup> | Postcentral_R | Temporal_Mid_L | Occipital_Mid_L | Cerebelm_Crus1_R |
| 9<sup>th</sup> | Cerebelm_Crus1_R | Parietal_Sup_L | Postcentral_L | Precuneus_L |
| 10<sup>th</sup> | Frontal_Sup_R | Temporal_Mid_R | Precuneus_L | Precentral_R |

**Figure 2.**
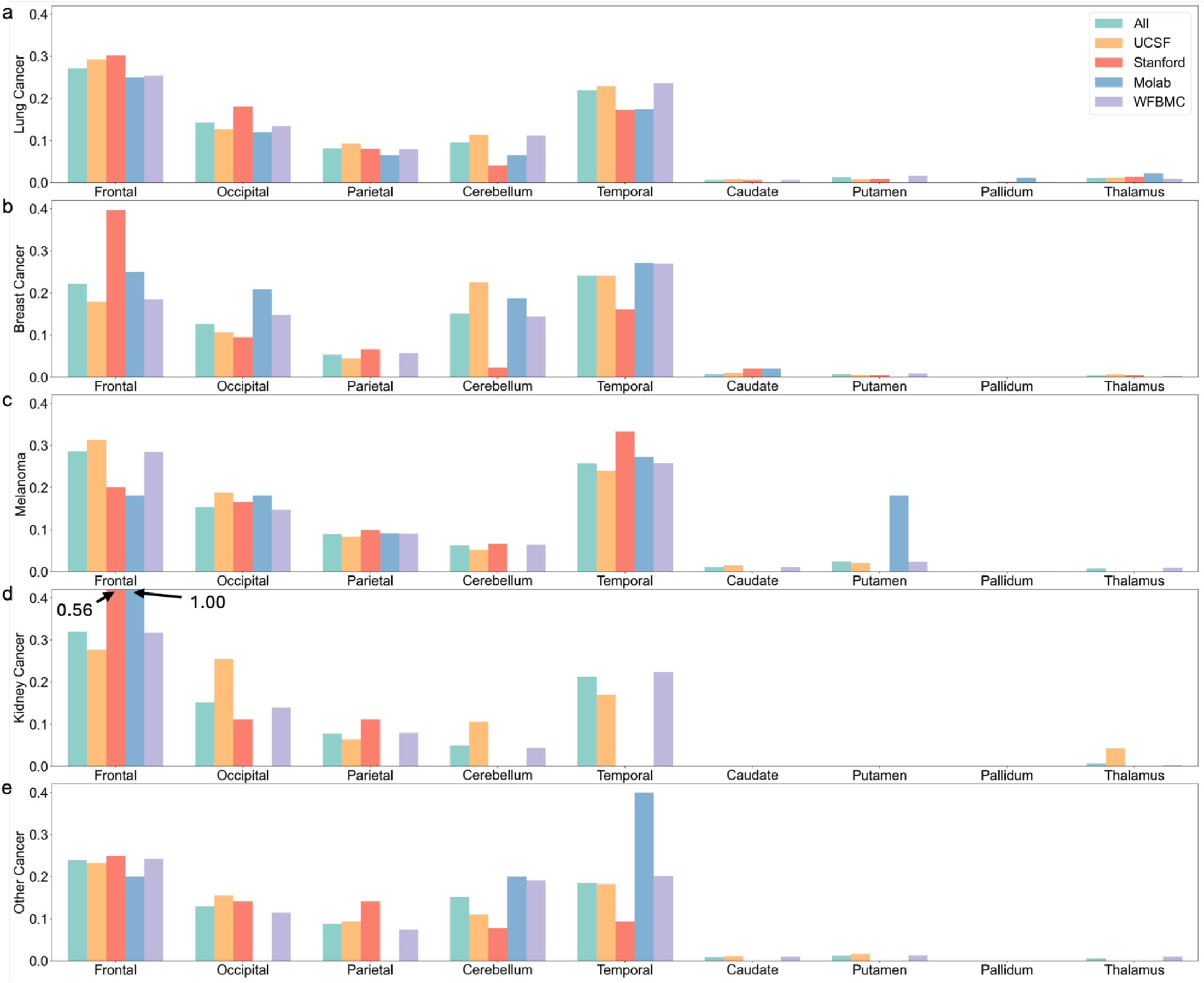
Frequency of different types of BM centroid locating in each region in the AAL Group atlas.

Figures 3(a) and 4(a) illustrate the distribution of lung cancer BM centroids across the first and second levels of Artery atlas. According to Figure 3(a), areas such as Anterior Cerebral Artery, Frontal pars of Middle Cerebral Artery, Parietal pars of Middle Cerebral Artery, Temporal pars of Middle Cerebral Artery, Occipital pars of Posterior Cerebral Artery, and Inferior Cerebellar are the most common sites for lung cancer BMs. At the second level of the Artery atlas, the Middle Cerebral Artery is the most frequently affected site.

**Figure 3.**
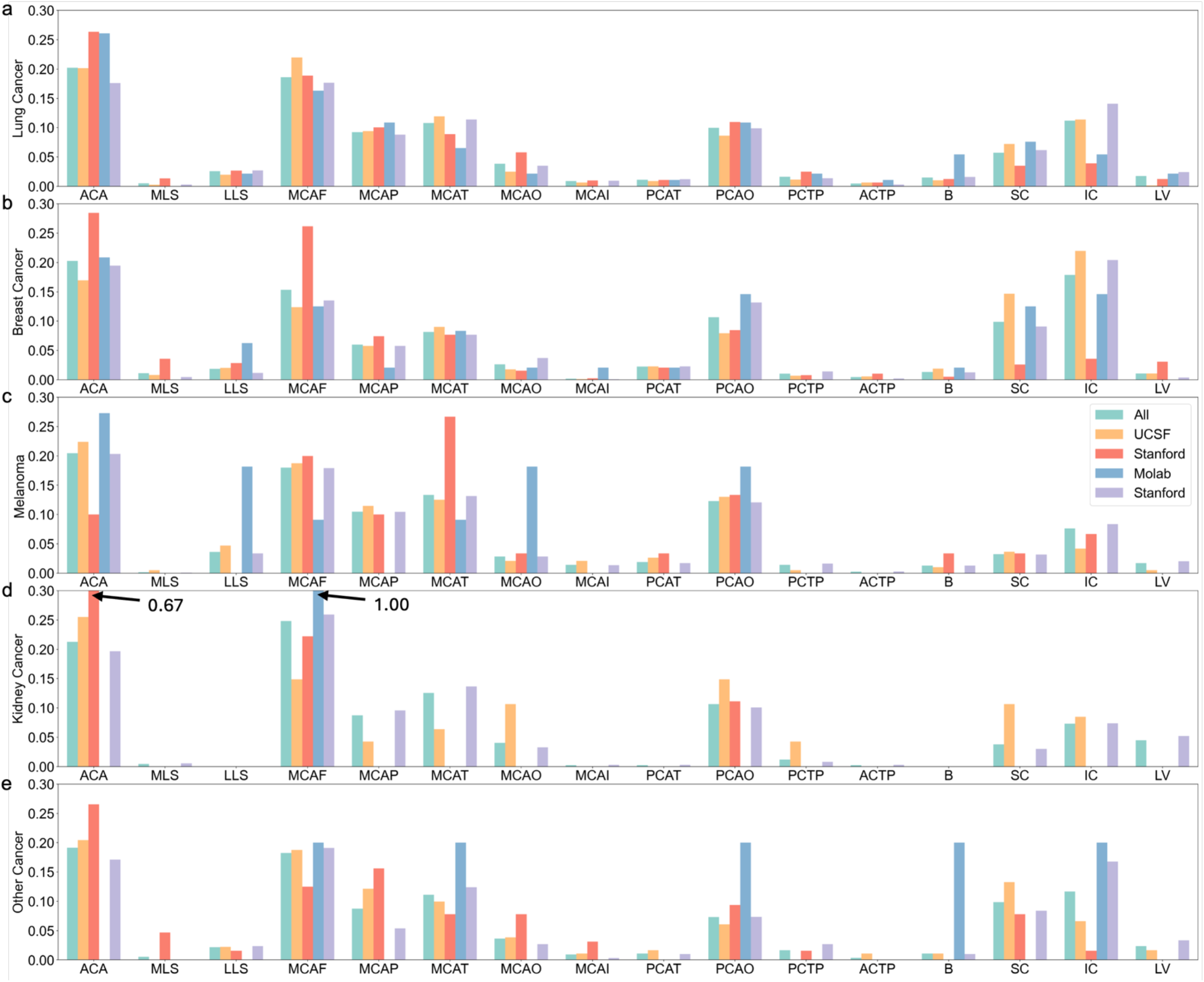
Frequency of different types of BM centroid locating in each region in the first level of Artery atlas. ACA: Anterior Cerebral Artery. MLS: Medial Lenticulostriate. LLS: Lateral Lenticulostriate. MCAF: Frontal pars of Middle Cerebral Artery. MCAP: Parietal pars of Middle Cerebral Artery. MCAT: Temporal pars of Middle Cerebral Artery. MCAO: Occipital pars of Middle Cerebral Artery. MCAI: Insular pars of Middle Cerebral Artery. PCAT: Temporal pars of Posterior Cerebral Artery. PCAO: Occipital pars of Posterior Cerebral Artery. PCTP: Posterior Choroidal and Thalamoperfurators. ACTP: Anterior Choroidal and Thalamoperfurators. B: Basilar. SC: Superior Cerebellar. IC: Inferior Cerebellar. LV: Lateral Ventricle.

**Figure 4.**
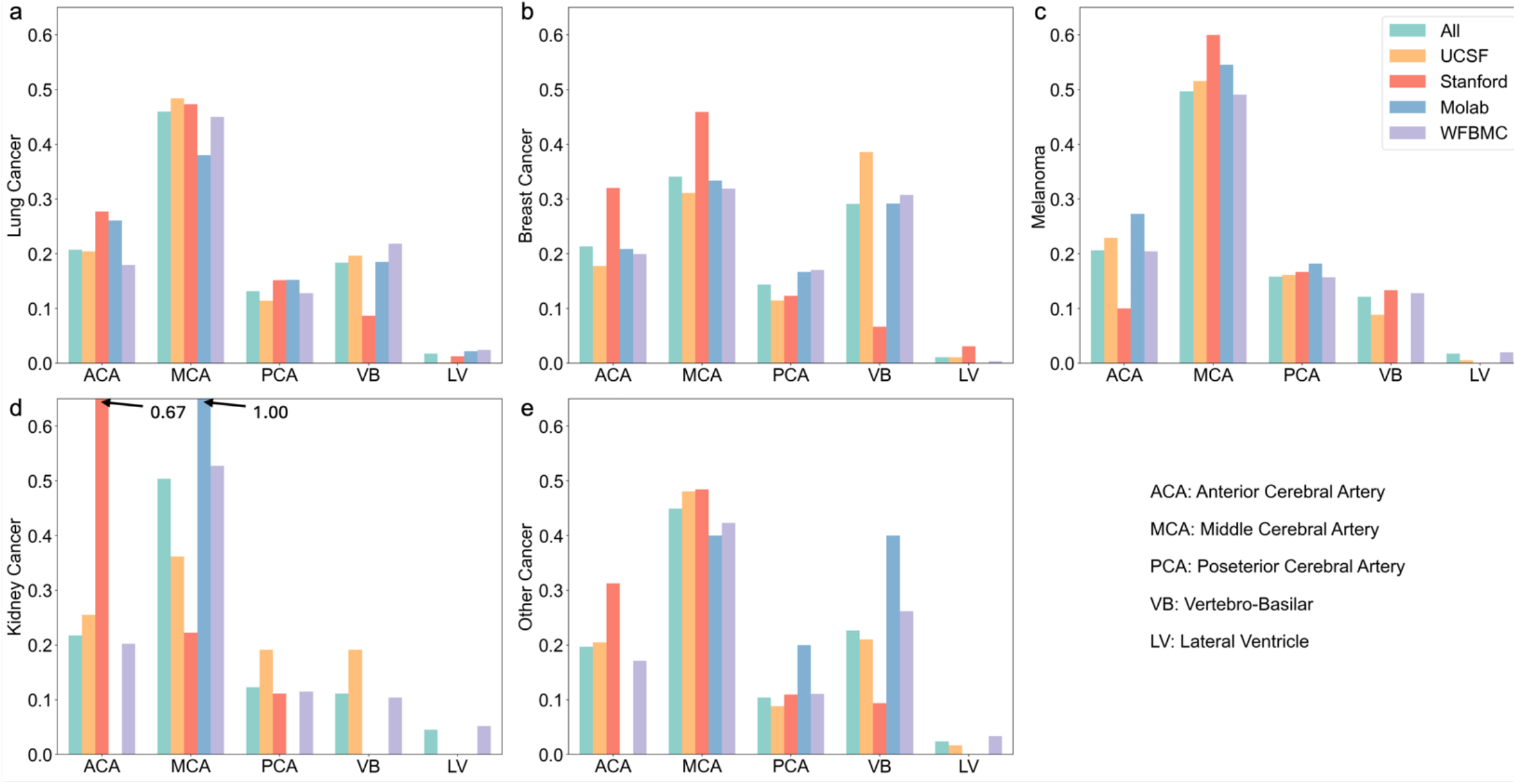
Frequency of different types of BM centroid locating in each region in the second level of Artery atlas.

### Breast cancer BM distribution

A total of 2,236 breast cancer BMs were investigated, accounting for 24% of all BMs involved in this study. Figure 5 shows the comparison of breast cancer BM frequency distribution maps from different datasets. It can be found that cerebellum, occipital, and temporal lobes are the most likely sites based on UCSF and WFBMC datasets, while frontal lobes are favored by UCSF and Stanford datasets.

**Figure 5.**
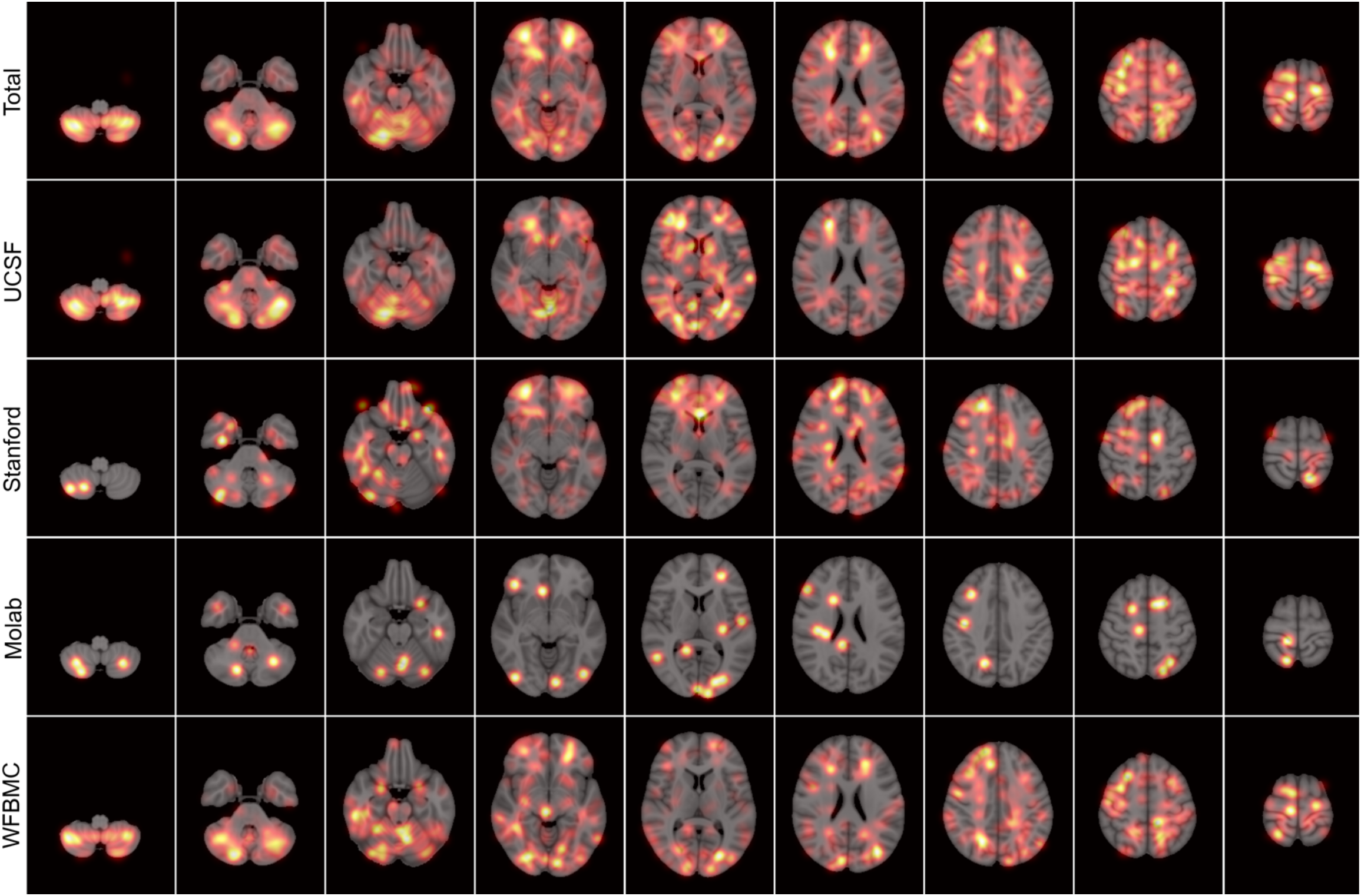
Breast cancer BM frequency distribution maps.

Like lung cancer BMs, we analyzed the distribution of breast cancer BM centroids using the AAL and AAL Group atlases. Figure 2(b) illustrates the frequency of lung cancer BM centroids in specific regions of the AAL Group atlas, while Table 3 identifies the ten most common AAL regions where breast cancer BMs were observed. The frontal and temporal regions emerge as the most prevalent across all datasets, with the Molab dataset showing a notable occurrence of BMs in the occipital region, and UCSF and Molab datasets showing a notable occurrence of BMs in the cerebellum. According to Table 3, there are multiple AAL regions locating in the cerebellum. Specifically, areas such as the Right Middle Frontal Gyrus, Left Cerebellum (Crus 1), and Right Cerebellum (Crus 1) appear in three out of four dataset results. Other regions, including the Left Cerebellum (Crus 2), Right Cerebellum (Crus 2), Right Cerebellum (Lobule 6), Left Cerebellum (Lobule 8), and Right Cerebellum (Lobule 8), were found in two of the four dataset results.

**Table 3.** The ten most common AAL regions for breast cancer BMs.

|  | UCSF | Stanford | Molab | WFBMC |
| --- | --- | --- | --- | --- |
| 1<sup>st</sup> | Cerebelum_6_R | Frontal_Mid_R | Cerebelum_Crus1_R | Cerebelum_Crus1_R |
| 2<sup>nd</sup> | Cerebelum_Crus1_R | Frontal_Sup_R | Cerebelum_Crus1_L | Frontal_Mid_R |
| 3<sup>rd</sup> | Cerebelum_8_R | Frontal_Mid_Orb_R | Parietal_Sup_L | Cerebelum_Crus2_R |
| 4<sup>th</sup> | Cerebelum_8_L | Frontal_Mid_L | Occipital_Sup_L | Cerebelum_8_L |
| 5<sup>th</sup> | Cerebelum_Crus2_L | Frontal_Sup_Medial_R | Vermis_6 | Cerebelum_Crus1_L |
| 6<sup>th</sup> | Cerebelum_Crus1_L | Frontal_Inf_Orb_R | Calcarine_L | Occipital_Mid_L |
| 7<sup>th</sup> | Cerebelum_Crus2_R | Frontal_Sup_Orb_R | Rectus_R | Cerebelum_8_R |
| 8<sup>th</sup> | Frontal_Mid_R | Fusiform_R | Supp_Motor_Area_R | Cerebelum_Crus2_L |
| 9<sup>th</sup> | Cerebelum_6_L | Angular_R | Temporal_Inf_L | Precentral_R |
| 10<sup>th</sup> | Temporal_Inf_R | Frontal_Sup_Orb_L | Temporal_Pole_Mid_R | Cerebelum_6_R |

Figures 3(b) and 4(b) depict the distribution of breast cancer BM centroids at both the first and second levels of Artery atlas. As shown in Figure 3(b), regions like the Anterior Cerebral Artery, Frontal pars of Middle Cerebral Artery, Occipital pars of Posterior Cerebral Artery, Superior Cerebellar, and Inferior Cerebellar are the most frequent sites for breast cancer BMs. At the second level of the Artery atlas, the Middle Cerebral Artery remains the most affected area. All datasets, except the Stanford dataset, indicate that the Vertebro-Basilar region is also a common site for breast cancer BMs. The Stanford dataset particularly highlights the Anterior Cerebral Artery as a key site region breast cancer BMs.

### Melanoma BM distribution

This study included 1,334 melanoma BMs, representing 14% of all BMs examined. Due to the limited number of melanoma BMs in the Molab dataset (only 11 cases), it is challenging to derive meaningful statistical results.

Consequently, this subsection focuses solely on the analysis from the other three datasets. Figure 6 presents frequency distribution maps of melanoma BMs. Unlike lung cancer and breast cancer BMs, melanoma BMs are more evenly distributed, with a slight preference for the occipital, temporal, and frontal lobes.

**Figure 6.**
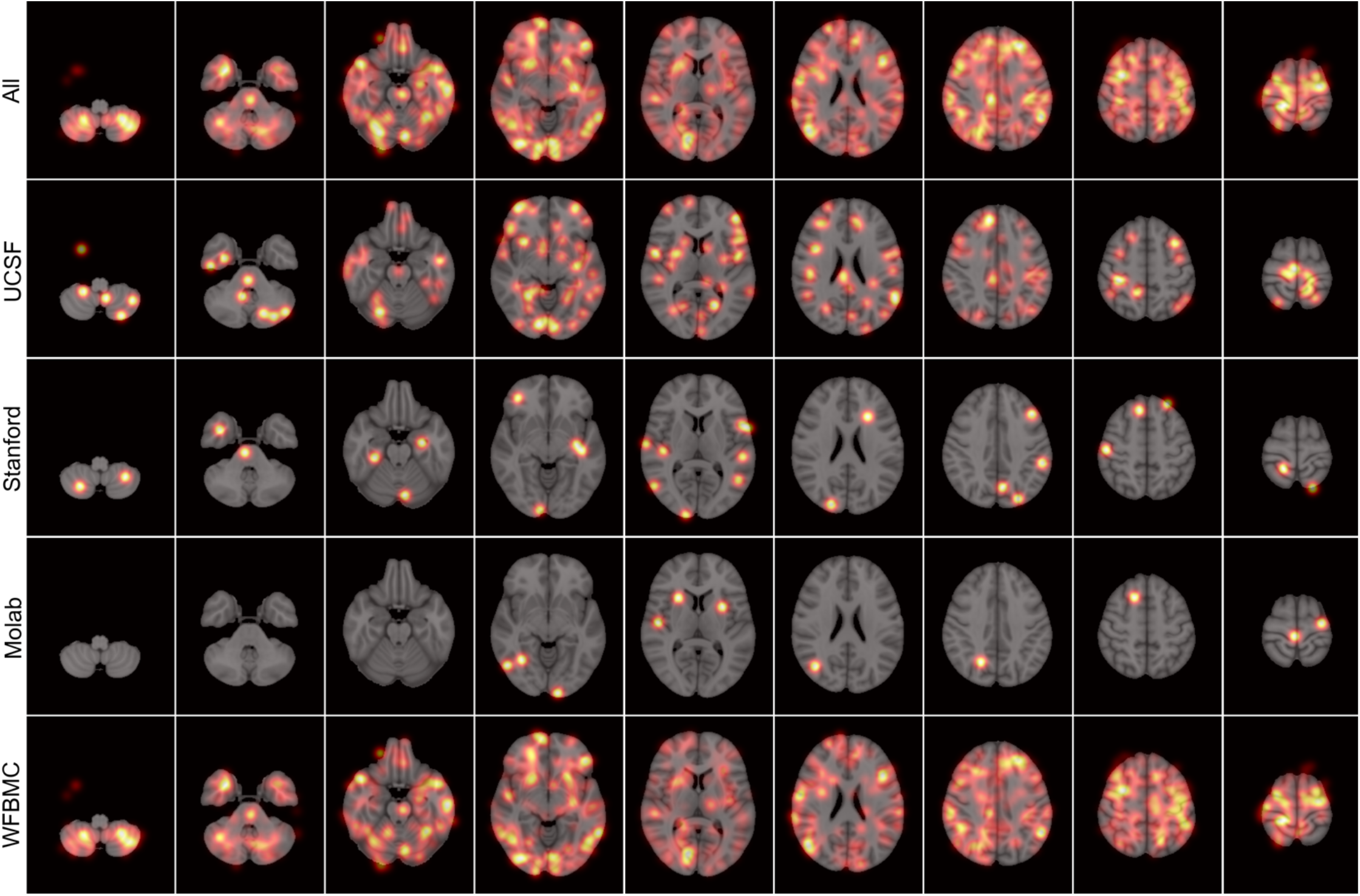
Melanoma BM frequency distribution maps.

Figure 2(c) illustrates the distribution of melanoma BMs in the AAL Group atlas. Consistent with Figure 6, which highlights the occipital, temporal, and frontal lobes as the most likely sites for melanoma BMs. Table 4 lists the ten most common AAL regions for melanoma BMs across the UCSF, Stanford, and WFBMC datasets. Several regions in the frontal, occipital, and temporal lobes are common to multiple datasets. Notably, the Left Middle Temporal Gyrus in the temporal lobe appears in all three datasets. Other AAL regions, such as the Right Middle Frontal Gyrus (frontal lobe), Right Postcentral Gyrus (occipital lobe), Right Superior Temporal Gyrus, and Right Middle Temporal Gyrus (temporal lobe), are found in two of the three datasets. Figures 3(c) and 4(c) demonstrate melanoma BM distribution on the Artery atlas. The Middle Cerebral Artery region is identified as the most frequent site for melanoma BMs across all datasets.

**Table 4.** The ten most common AAL regions for melanoma BMs.

|  | UCSF | Stanford | WFBMC |
| --- | --- | --- | --- |
| 1<sup>st</sup> | Frontal_Mid_R | Precuneus_L | Frontal_Mid_R |
| 2<sup>nd</sup> | Temporal_Mid_L | Postcentral_R | Frontal_Sup_R |
| 3<sup>rd</sup> | Calcarine_L | Cerebelum_Crus1_L | Postcentral_R |
| 4<sup>th</sup> | Lingual_R | Temporal_Mid_R | Calcarine_R |
| 5<sup>th</sup> | Parietal_Inf_L | Temporal_Mid_L | Precentral_L |
| 6<sup>th</sup> | Cingulum_Mid_R | Temporal_Sup_R | Temporal_Inf_L |
| 7<sup>th</sup> | Frontal_Inf_Tri_L | Temporal_Sup_L | Temporal_Mid_R |
| 8<sup>th</sup> | Frontal_Sup_Medial_R | Heschl_R | Frontal_Mid_L |
| 9<sup>th</sup> | Temporal_Sup_R | Fusiform_R | Supp_Motor_Area_L |
| 10<sup>th</sup> | Postcentral_L | Occipital_Sup_R | Temporal_Mid_L |

### Kidney cancer BM distribution

Approximately 4% of all BMs involved in this study are from kidney cancer. Due to the limited number of kidney BMs in the Stanford and Molab datasets, it was challenging to derive meaningful statistical results. Consequently, this subsection focuses solely on the analysis from the remaining two datasets. Figure 7 presents frequency distribution maps based on 423 kidney cancer BMs, showing that the frontal and temporal lobes are the most likely sites for these metastases.

**Figure 7.**
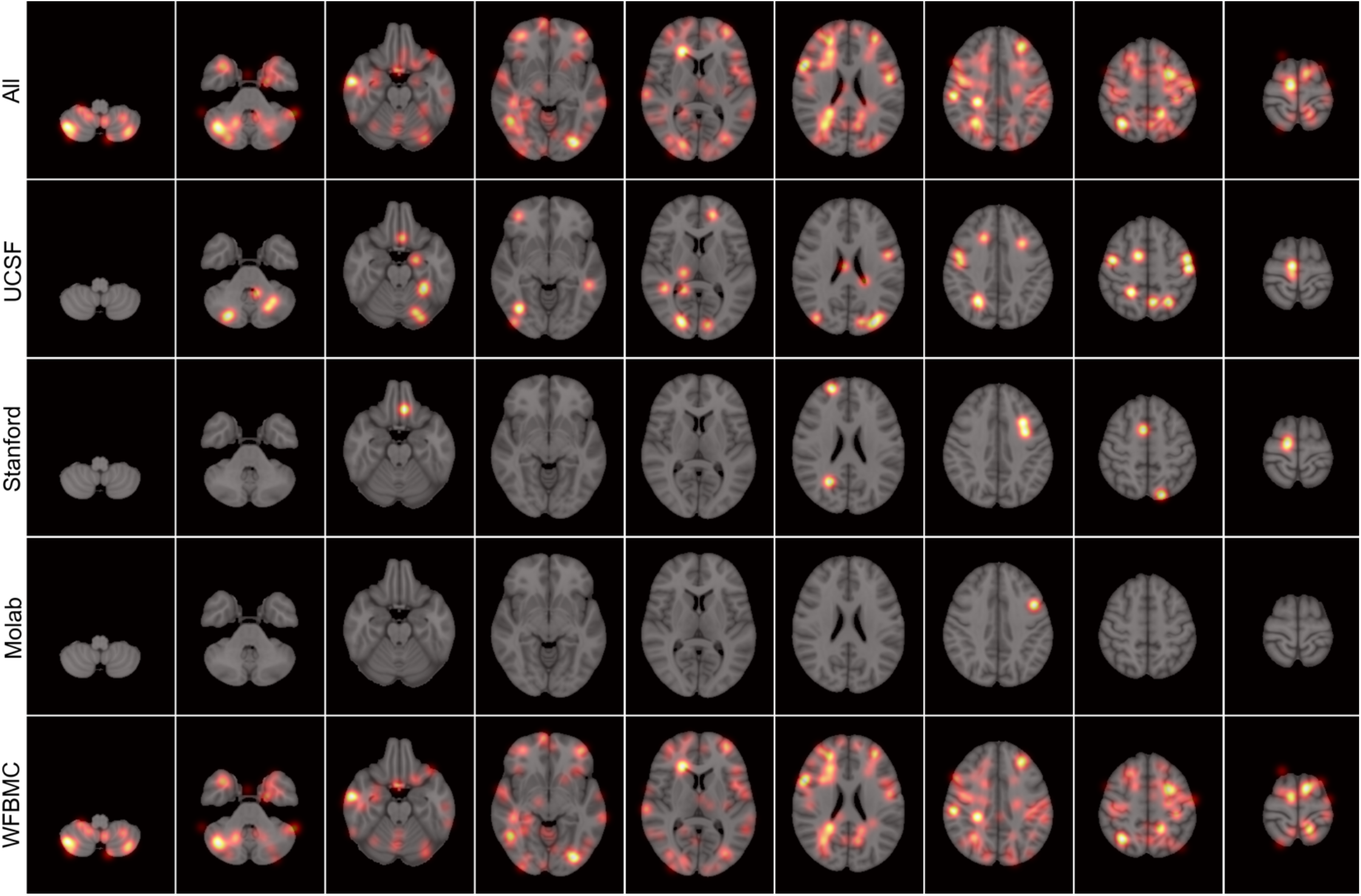
Kidney cancer BM frequency distribution maps.

Figure 2(d) illustrates the distribution of kidney BMs in the AAL Group atlas. Consistent with Figure 7, the frontal and temporal lobes are hightlighted as the most probable sites for kidney BMs. Table 5 lists the ten most common AAL regions for kidney BMs across the UCSF and WFBMC datasets. The Right Precentral Gyrus in the frontal lobe and Left Postcentral Gyrus in the occipital lobe appears in all two datasets. On the Artery atlas, the Anterior Cerebral Artery and Middle Cerebral Artery regions emerge as the most common sites for kidney cancer BMs.

**Table 5.** The ten most common AAL regions for kidney cancer BMs.

|  | UCSF | WFBMC |
| --- | --- | --- |
| 1<sup>st</sup> | Supp_Motor_Area_R | Frontal_Mid_L |
| 2<sup>nd</sup> | Precentral_R | Temporal_Mid_R |
| 3<sup>rd</sup> | Cerebelum_6_L | Frontal_Sup_L |
| 4<sup>th</sup> | Occipital_Mid_L | Frontal_Sup_R |
| 5<sup>th</sup> | Thalamus_R | Precuneus_L |
| 6<sup>th</sup> | Occipital_Inf_R | Precentral_L |
| 7<sup>th</sup> | Cerebelum_Crus1_L | Postcentral_L |
| 8<sup>th</sup> | Frontal_Sup_Medial_L | Precentral_R |
| 9<sup>th</sup> | Postcentral_L | Frontal_Mid_R |
| 10<sup>th</sup> | Temporal_Sup_R | Frontal_Inf_Oper_R |

**Table 6.** MANOVA results on BM centroids across four datasets.

|  |  | Value | F-value | p-value |
| --- | --- | --- | --- | --- |
| Lung | Wilks' lambda | 0.9837 | 8.9422 | <0.001 |
|  | Pillai's trace | 0.0163 | 8.9016 | <0.001 |
|  | Hotelling-Lawley trace | 0.0165 | 8.9736 | <0.001 |
|  | Roy's greatest root | 0.0153 | 24.8682 | <0.001 |
| Breast | Wilks' lambda | 0.8942 | 28.3800 | <0.001 |
|  | Pillai's trace | 0.1067 | 27.4465 | <0.001 |
|  | Hotelling-Lawley trace | 0.1174 | 29.0986 | <0.001 |
|  | Roy's greatest root | 0.1087 | 80.9248 | <0.001 |
| Melanoma | Wilks' lambda | 0.9937 | 0.9299 | 0.4976 |
|  | Pillai's trace | 0.0063 | 0.9305 | 0.4970 |
|  | Hotelling-Lawley trace | 0.0063 | 0.9296 | 0.4979 |
|  | Roy's greatest root | 0.0042 | 1.8654 | 0.1336 |
| Kidney | Wilks' lambda | 0.9585 | 1.9835 | 0.0381 |
|  | Pillai's trace | 0.0420 | 1.9815 | 0.0381 |
|  | Hotelling-Lawley trace | 0.0429 | 1.9845 | 0.0386 |
|  | Roy's greatest root | 0.0290 | 4.0536 | 0.0074 |

### Other BM distribution

In this study, all cancer types besides lung, breast, melanoma, and kidney were categorized as “other”, with a total of 548 BMs, representing around 6% of all BMs. Figure 8 presents frequency distribution maps for these metastases, showing that areas such as the cerebellum, frontal, temporal, occipital, and parietal lobes are all potential sites for BMs. Since these BMs originate from various smaller categories, including rectal, head and neck, and prostate cancers, we provide an overall frequency distribution. Due to the limited number of cases in each subtype, we did not analyze the distribution of individual subtypes.

**Figure 8.**
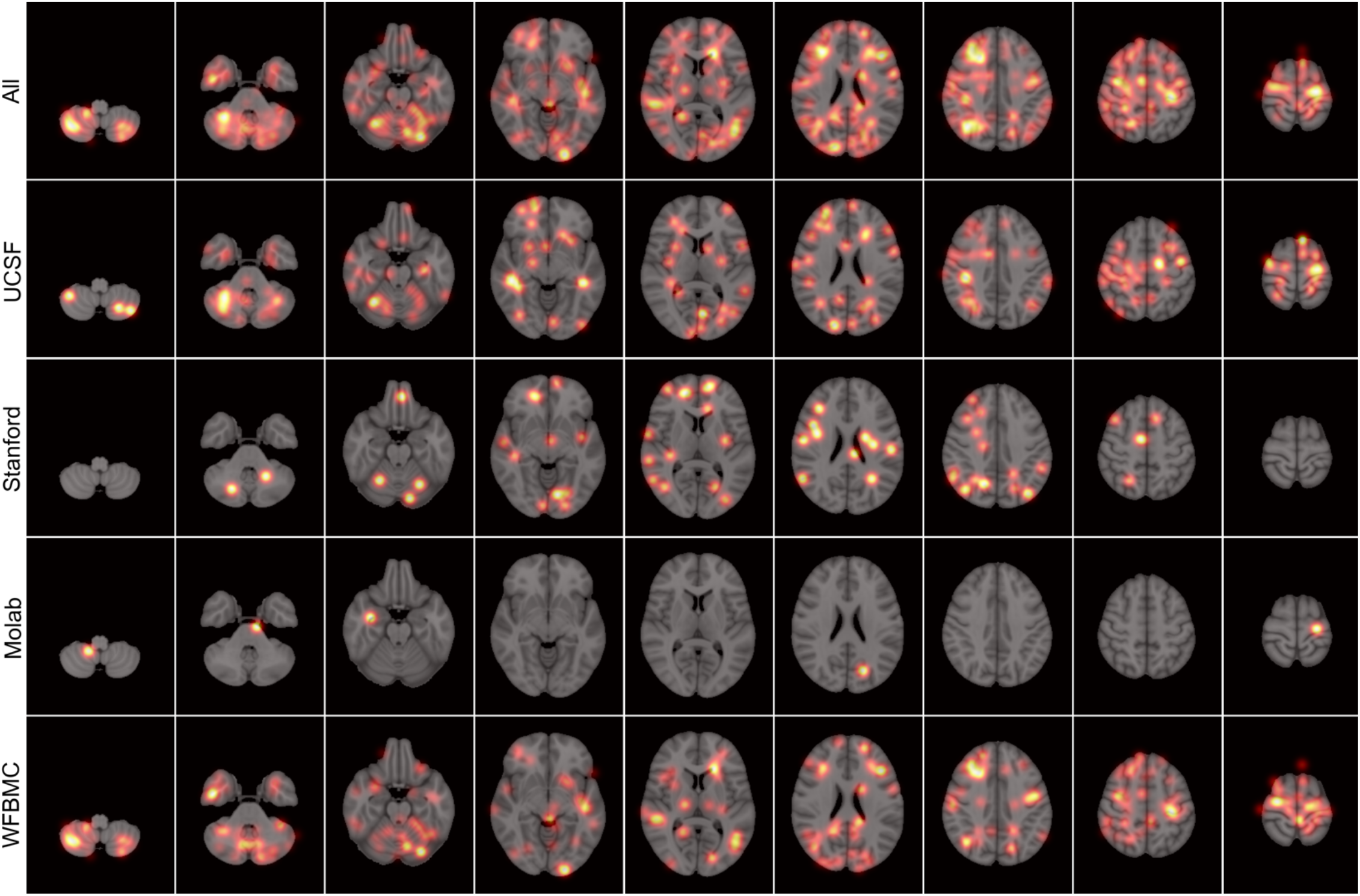
Other BM frequency distribution maps.

### BM volumes analysis

This study also examined the volume of BMs for each cancer type. According to Figure 9, lung, breast, and other BMs tend to have the highest counts in the smaller volume group (less than 0.1 cm^3^), with most of these BMs measuring less than 0.5 cm^3^. In contrast, melanoma and kidney cancer BMs are slightly larger, with the majority falling into the 0.1 - 0.5 cm^3^ range. Among the lung, breast, melanoma, and kidney cancer BMs, kidney cancer BMs display the most even distribution in terms of volume.

**Figure 9.**
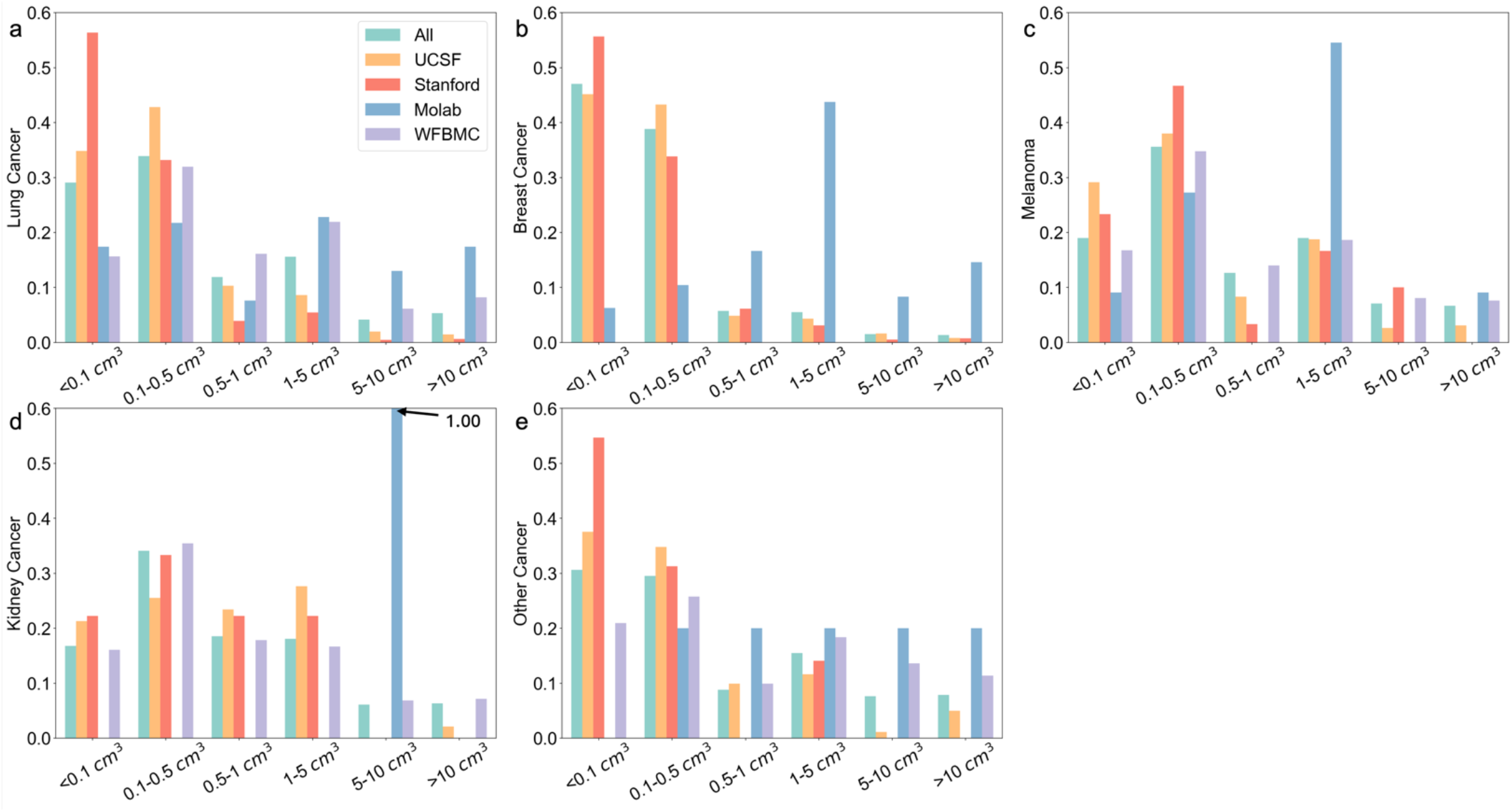
Volume frequency distribution of different BM types.

### BM distribution similarity analysis across different datasets

It is crucial to determine if there are significant differences in BM distribution across various datasets, as this helps us understand the consistency of BM distribution patterns. To assess this, we employed Multivariate Analysis of Variance (MANOVA) to investigate the consistency of BM distribution across different datasets. For each dataset, tumor centroid position information was extracted and used for the analysis.

The breast cancer BM distribution shows similar results to lung cancer BMs, with significant differences in tumor centroid distributions across datasets. However, the actual differences in centroid positions between datasets are not large in magnitude. For melanoma BMs, the MANOVA results indicate no significant differences in tumor centroid distributions across datasets. In contrast, kidney BMs show statistically significant differences in tumor centroid distributions across datasets, though the actual magnitude of these differences is minimal. The p-values confirm that the differences are not due to random chance, but the close-to-1 Wilks’ Lambda values suggest that the tumor centroid distributions across datasets remain largely similar.

## Discussion

In this study, we investigated the distribution of various types of BMs using four datasets. The distribution of lung, breast, melanoma, and kidney cancer BMs were analyzed and compared with findings from existing studies. We compared the distribution of BM across different datasets. While we observed significant differences in tumor centroid distributions across datasets, the actual differences in centroid positions were not large in magnitude. As a result, these differences are unlikely to significantly impact the overall investigation results.

Our investigation revealed that lung cancer BMs are most commonly located in the cerebellum, as well as the frontal, temporal, occipital, and parietal lobes, consistent with findings from previous studies^11-15^. Breast cancer BMs frequently occur in the cerebellum and the frontal, occipital, and temporal lobes, which aligns with prior research^11,12,15,17^. For melanoma BMs, the occipital, temporal, and frontal lobes are the most common regions, similar to findings by Neman *et al*. and Schroeder *et al*.^2,15^. Although studies on kidney cancer BM distribution are limited, some suggest that kidney cancer BMs may occur in regions like the brainstem and deep white matter^9,15^. In our analysis of 423 kidney cancer BMs, we found that the frontal and temporal lobes are the most likely sites for these metastases.

We further explored the potential relationship between BM volume and BM type. Our findings indicate that lung and breast cancer BMs tend to be smaller, with the majority having a volume of less than 0.5 cm^3^. In contrast, melanoma and kidney cancer BMs have a more evenly distributed volume range, with a higher likelihood of larger tumors exceeding 1 cm^3^ in size.

Given the heterogeneity in intracranial distribution and tumor volume across different BM types, it is possible to develop algorithms that identify BM types based on structural MRI. This could accelerate the BM diagnosis process. In approximately 10% of cases, brain metastatic disease is the initial cancer presentation^34^. Additionally, in up to 15% of BM patients, the primary tumor remains unidentified^35^. Developing fast BM identification algorithms would benefit these patients by enabling timely management of the extracranial tumor and improving prognosis. There are several studies have aimed to create algorithms to identify the primary cancer type of BMs using brain MRI scans and the inherent BM distribution patterns^36-39^. Our study will support these efforts and contribute to more accurate BM type identification in the future.

## Data Availability

All open-access datasets used in this study are available online. Please go to official websites for data access.

